# Diagnostic Performance of Claude 3 from Patient History and Key Images in Diagnosis Please Cases

**DOI:** 10.1101/2024.04.11.24305622

**Authors:** Ryo Kurokawa, Yuji Ohizumi, Jun Kanzawa, Mariko Kurokawa, Takao Kiguchi, Wataru Gonoi, Osamu Abe

## Abstract

**Backgrounds:** Large language artificial intelligence models have showed its diagnostic performance based solely on textual information from clinical history and imaging findings. However, the extent of their performance when utilizing radiological images and providing differential diagnoses has yet to be investigated.

**Purpose:** We employed the latest version of Claude 3, Opus, released on March 4, 2024, to investigate its diagnostic performance in answering Radiology’s Diagnosis Please quiz questions under three conditions: (1) when provided with clinical history alone; (2) when given clinical history along with imaging findings; and (3) when supplied with clinical history and key images.

Furthermore, we evaluated the diagnostic performance of the model when instructed to list differential diagnoses.

**Materials and Methods:** Claude 3 Opus was tasked with listing the primary diagnosis and two differential diagnoses for 322 quiz questions from Radiology’s “Diagnosis Please” cases, which included cases 1 to 322, published from 1998 to 2023. The analyses were carried out under the following input conditions:

Condition 1: Submitter-provided clinical history (text) alone

Condition 2: Submitter-provided clinical history and imaging findings (text) Condition 3: Submitter-provided clinical history (text) and key images (PDF files)

We applied McNemar’s tests to evaluate differences in correct response rates for primary diagnoses across Conditions 1, 2, and 3.

**Results:** The correct primary diagnoses rates were 62/322 (19.3%), 178/322 (55.3%), and 93/322 (28.8%) for Conditions 1, 2, and 3, respectively. Additionally, Claude 3 Opus accurately provided the correct answer as a differential diagnosis in up to 22/322 (6.8%) of cases. There were statistically significant differences in correct response rates for primary diagnoses between all combinations of Conditions 1, 2, and 3 (p<0.001).

**Conclusion:** Claude 3 Opus demonstrated significantly improved diagnostic performance by inputting key images in addition to clinical history. The ability to list important differential diagnoses was also confirmed.

**Key Results:**

- This study investigated Claude 3 Opus’s performance in Radiology Diagnosis Please Cases using clinical history, key images, and imaging findings.
- Key images or imaging findings inputs significantly improved correct primary diagnoses from 19.3% to 28.8% or 55.5%, respectively.
- By having two additional differential diagnoses presented, total correct responses improved by 3.1–6.8%.

**Summary statement:** Large language AI model Claude 3 Opus demonstrated significantly improved diagnostic accuracy by adding key images with clinical history compared with clinical history alone.

## Introduction

Large language artificial intelligence (AI) models are designed to understand and generate human-like text based on the input they receive, and have demonstrated remarkable capabilities across various domains[1]. In the field of diagnostic imaging, Ueda et al.[2] conducted a study using Chat GPT’s GPT-4 model[3] and reported that it correctly answered 54% (170 of 313) of Radiology’s Diagnosis Please quiz questions based solely on textual information from clinical history and imaging findings. This highlights the potential use of language AI in diagnostic imaging. However, the diagnostic accuracy of the AI models using radiological images remains unclear. Moreover, the ability to provide a list of highly probable differential diagnoses and their respective accuracies, which is crucial in daily clinical practice, has not been previously investigated using AI models. Claude 3[4], a large-language AI model developed by Anthropic (California, United States), possesses the ability to read and analyze not only input text but also image data. In this study, we employed the latest version of Claude 3, Opus, released on March 4, 2024, to examine its diagnostic performance in answering Radiology’s Diagnosis Please quiz questions under three conditions: (1) when provided with clinical history alone; (2) when given clinical history along with imaging findings; and (3) when supplied with clinical history and key images. Furthermore, we evaluated the diagnostic performance of the model when instructed to list differential diagnoses.

## Materials and Methods

An overview of this study is presented in Figure (**Figure**). We used Claude 3 Opus (Anthropic, California, United States, released on 4th Mar 2024, accessed on 15th Mar 2024, https://claude.ai/) to list the primary diagnosis and two differential diagnoses for 322 quiz questions from Radiology’s Diagnosis (https://dxp.rsna.org/), spanning from Cases 1 to 322, published between 1998 and 2023. Analyses were conducted under the following three input data conditions:

**Figure.**
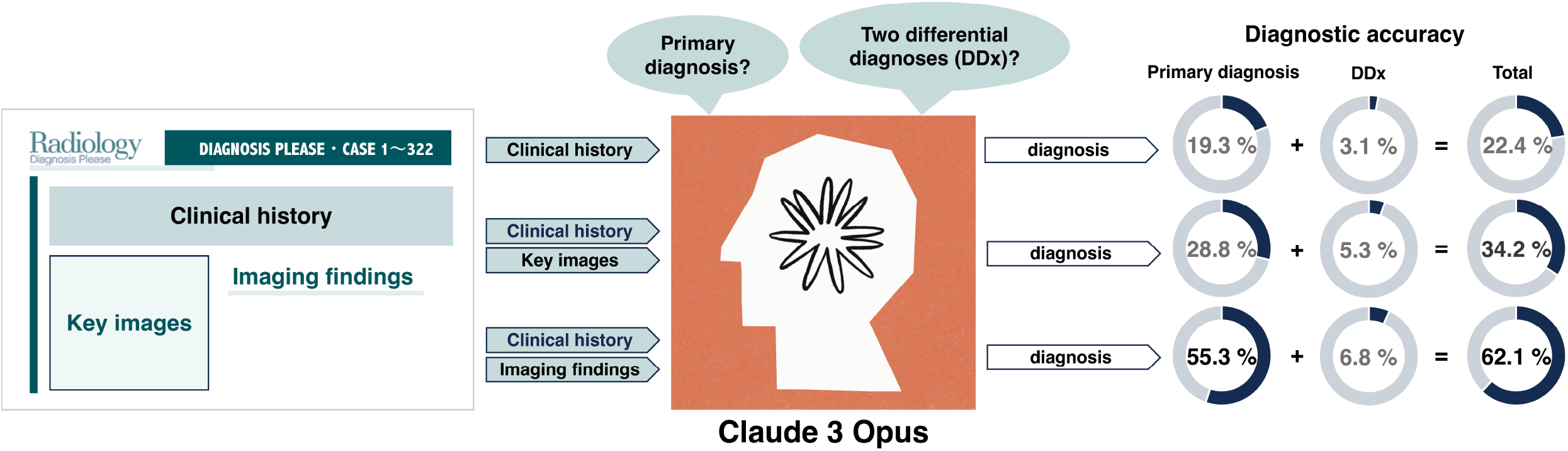
Overview of this study. Diagnostic Performance of Claude 3 from Patient History and Key Images in Diagnosis Please Cases. • This study investigated Claude 3 Opus’s performance in Radiology Diagnosis Please Cases using clinical history, key images, and imaging findings. • Key images or imaging findings inputs significantly improved correct primary diagnoses from 19.3% to 28.8% or 55.5%, respectively. • By having two additional differential diagnoses presented, total correct responses improved by 3.1-6.8%.

Condition 1: Submitter-provided clinical history (text) alone

Each condition was examined using separate threads. When extracting the submitter-identified imaging findings, two trainee radiologists and one board-certified diagnostic radiologist with 11 years of experience meticulously removed sentences containing answers to ensure the integrity of the analysis. The accuracy of the primary diagnosis and the two differential diagnoses generated by Claude 3 Opus were determined by consensus among three board-certified diagnostic radiologists with 8, 11, and 19 years of experience. Ethical approval was not required because this study exclusively utilized published articles. McNemar’s tests were used to assess the difference in correct response rates for the primary diagnoses under Conditions 1, 2, and 3. Two-sided P values <0.05 were considered statistically significant. Statistical analyses were performed using the R software (version 4.1.1; R Foundation for Statistical Computing, Vienna, Austria).

## Results

The results are summarized in Table (**Table**). The primary diagnoses were correct in 62/322 (19.3%), 178/322 (55.3%), and 93/322 (28.8%) cases under conditions 1, 2, and 3, respectively. Claude 3 Opus provided the correct answer as a differential diagnosis in up to 22/322 (6.8%) cases. There were significant differences in the correct response rates for primary diagnoses between any combination of conditions 1, 2, and 3 (ps<0.001).

**Table.** Summary of the results.

**Table. Summary of the results**
| Input data | Correct primary<br>diagnosis | Correct differential<br>diagnoses | Total |
| --- | --- | --- | --- |
| Condition 1: Clinical history alone | 62/322 (19.3%) | 10/322 (3.1%) | 72/322<br>(22.4%) |
| Condition 2: Clinical history and<br>imaging findings | 178/322 (55.3%) | 22/322 (6.8%) | 200/322<br>(62.1%) |
| Condition 3: Clinical history and<br>key images | 93/322 (28.8%) | 17/322 (5.3%) | 110/322<br>(34.2%) |

## Discussion

In this study, we evaluated the diagnostic performance of the language AI model Claude 3 Opus in answering Radiology’s Diagnosis Please quiz questions based on three types of input information: clinical history alone, clinical history accompanied by imaging findings, and clinical history coupled with key images.

Ueda et al. [2] demonstrated that using the Chat GPT GPT-4 model, a large language AI model, with the input of text information from clinical history and imaging findings, Chat GPT correctly answered 54% of questions. Similarly, Li et al. [5] showed a 17% improvement in diagnostic accuracy when using the GPT-4 model compared to the GPT-3.5 model of Chat GPT when inputting text information from clinical history and imaging findings. Large language AI models continue to evolve, enabling them to read the image information provided by users.

However, the diagnostic performances of AI models that use radiological images in conjunction with textual information have not yet been investigated.

The results of this study demonstrated that Claude 3 Opus could present the correct diagnosis in 28.8% (93/322) of cases by inputting key images in addition to the clinical history. This shows a 9.5% increase with a statistically significant difference in accuracy compared with providing clinical history alone, potentially reflecting the imaging diagnostic performance of AI models. However, it was inferior to the accuracy achieved with clinical history and submitter-provided imaging findings (28.8% vs 55.3%; respectively, with a significant difference in accuracy of 26.5%). The 55.3% (178/322) accuracy of primary diagnoses obtained by Claude 3 Opus from clinical history plus imaging findings was nearly equivalent to the 54% shown by Ueda et al. using the GPT-4 model.

Owing to real-world complexity, there may be more than one possible diagnosis in daily clinical practice, necessitating the reporting of potential differential diagnoses. In this study, the AI model presented two differential diagnoses, in addition to the primary diagnosis. It succeeded in presenting the correct answer as a differential diagnosis in up to 6.8% (22/322) of cases, resulting in an accuracy of up to 62.1 % (200/322) when the clinical history and imaging findings were input.

The results of this study suggest that large language AI models, in their current state, may be more effectively utilized as a supportive tool for radiologists in formulating differential diagnoses based on accurate image interpretation and clinical history, rather than as a standalone solution for replacing radiologists in the image interpretation process. Although these AI models demonstrate a certain level of usefulness in assisting radiologists, their ability to interpret image findings solely from the images themselves and provide meaningful contributions to diagnosis still requires improvement and is still in the early stages of development. However, as the performance of large language AI models continues to advance and innovations in the provided information emerge, there is potential for further improvements in their diagnostic capabilities. To fully realize this potential, additional research and validation are necessary to explore the evolving role of AI models in the field of diagnostic imaging.

## Data Availability

All data produced are available online at https://pubs.rsna.org/journal/radiology.

## Abbreviations

AI: artificial intelligence

## Notes

### Competing Interest Statement

The authors have declared no competing interest.

### Funding Statement

This study did not receive any funding

